# High *Coxiella burnetii* seroconversion rate in veterinary students, the Netherlands, 2006-2010

**DOI:** 10.1101/2020.06.10.20126185

**Authors:** Marit M.A. de Lange, Wim van der Hoek, Peter M. Schneeberger, Arno Swart, Dick J. J. Heederik, Barbara Schimmer, Inge M. Wouters

**Author notes:** **Corresponding author:** Marit M.A. de Lange, PhD, epidemiologist, National Institute for Public Health and the Environment, Centre for Infectious Disease Control Netherlands, Antonie van Leeuwenhoeklaan 9, 3721 MA, Bilthoven, the Netherlands.

## Abstract

We examined seroconversion rates by measuring IgG antibodies against *Coxiella burnetii* among two cohorts of veterinary students. During follow-up of 118 seronegative veterinary students, 23 students seroconverted. Although the clinical significance of the presence of antibodies is unknown, students should be informed about the potential risks of Q fever.

## Text

Q fever is caused by the bacterium *Coxiella burnetii* and can present as acute or chronic illness. However, around 60% of infected people remain asymptomatic *(1)*. In particular, livestock veterinarians are at increased risk of a *C. burnetii* infection *(2)*. Previously, a high seroprevalence, range 11-19%, among veterinary medicine students was reported *(3-5)*. However, the incidence of Q fever and associated risk factors during veterinary training are still unknown. Therefore, we performed a longitudinal study in veterinary students in the Netherlands following incoming, seronegative veterinary students during a maximum of 4-year study period and registered potential study and non-study related associated factors for seroconversion.

## The study

Veterinary medicine students who started in 2006 or 2008 (around 225 each year) at the Faculty of Veterinary Medicine of Utrecht University (FVMUU) were eligible for inclusion. This faculty provides the only veterinary medicine program in the Netherlands. Recruitment methods included informational class presentations and a mailed brochure. The Medical Ethical Commission of the University Medical Centre Utrecht approved the study protocol (no. 06/169). After obtaining written informed consent, a blood sample was collected at study start and participants completed a baseline questionnaire. From participants who started at the FVMUU in 2006 (cohort 2006), up to two additional blood samples were collected and follow-up questionnaires completed in 2008 and 2010. Students who started in 2008 (cohort 2008) provided only one follow-up blood sample and follow-up questionnaire, in 2010. Both the baseline and follow-up questionnaires included questions about animal contact, living situation, personal health situation, and smoking habits before and during the study period. The follow-up questionnaires also included questions about the study choices and study related courses.

Serum samples were tested for IgG antibodies against phase I and II of *C. burnetii*, using an indirect immunofluorescence assay (IFA) as previously described *(3)*. Those with IgG phase I or II antibodies ≥ 1:32 were classified as *C. burnetii* seropositive. Seroconversion was defined as a participant who was IgG seronegative at baseline and seropositive in one of the follow-up samples. Participants with an IgG phase I titer of ≥ 1:1024 had a serological indication for a chronic Q fever infection *(6)*.

All data were analyzed with SAS, version 9.4 (SAS Institute Inc., USA). First, differences in demographics and past animal exposure characteristics between seropositive and seronegative participants at baseline were determined with a Fisher exact test (numerical characteristics) or Kruskal Wallis test (categorical characteristics). To estimate seroconversion rate and possible associated factors for seroconversion during follow-up, data from seronegative participants with at least one follow-up sample were used. The univariable logistic regression analyses were performed with generalized estimating equations (GEE) models with an exchangeable correlation matrix. These models were used to take into account correlations between the repeated measurements of serostatus within the same subject *(7)*. Participants’ data were censured for the times after they were tested *C. burnetii* seropositive. The data from the two cohorts were analyzed together, because the data sets were too small to analyze them separately. The FVMUU starting year (cohort) and the number of years after the study start were always included as covariates in the model. Investigated characteristics were animal-related exposure outside the study, living situation, smoking habits, study duration, cohort, and chosen study direction; in total, we investigated 20 characteristics. Associations were considered significant at confidence level of α < 0.05. All univariable associated characteristics were highly interrelated (P<0.05 in Fisher exact test). Therefore, multivariable logistic GEE analysis was not possible.

At the beginning of their veterinary training 447 students were invited to participate in the study of which 131 participated, of whom 13 (10%) were *C. burnetii* IgG seropositive at baseline. Students who were seropositive at baseline were more likely to have ever lived on a farm (Table 1). Similarly, they had more contact with farm animals than students who were seronegative at baseline, but the difference was only statistically significant for contact with cows and poultry. No other significant differences were found between *C. burnetii* seronegative and seropositive students at baseline (Table 1).

**Table 1.** Baseline questionnaire characteristics of two cohorts of veterinary students (cohort 1 started in 2006, cohort 2 started in 2008) at the Faculty of Veterinary Medicine of Utrecht University, the Netherlands.

| Characteristic | Seronegative participants<br>at baseline in 2006 or<br>2008 N=118*<br>Median or no/N (%) | Seropositive<br>participants at baseline<br>in 2006 or 2008 N=13<br>Median or no/N (%) | p value |
| --- | --- | --- | --- |
| Age (years) | 19 | 18 | 0.24 |
| BMI | 21 | 21 | 0.87 |
| Gender |  |  |  |
| Male | 18/116 (16) | 3/13 (23) | 0.44 |
| Female | 98/116 (84) | 10/13 (77) |  |
| Smoking |  |  |  |
| No smoking | 109/115 (94) | 13/13 (100) | 1.00 |
| Past smoker | 3/115 (3) | 0/13 (0) |  |
| Current smoker | 3/115 (3) | 0/13 (0) |  |
| Grew up in a |  |  |  |
| Village (<15,000 inhabitants) | 44/116 (38) | 7/13 (54) | 0.45 |
| Town (15,000-80,000 inhabitants) | 47/116 (40) | 5/13 (38) |  |
| City (>80,000 inhabitants) | 25/116 (22) | 1/13 (8) |  |
| Ever lived on a farm |  |  |  |
| Yes | 11/116 (9) | 7/13 (54) | <0.01 |
| No | 105/116 (91) | 6/13 (46) |  |
| Regular contact with cows before start of FVMUU |  |  |  |
| Yes | 19/116 (16) | 9/13 (69) | <0.01 |
| No | 97/116 (84) | 4/13 (31) |  |
| Regular contact with goats before start of FVMUU |  |  |  |
| Yes | 18/116 (16) | 5/13 (38) | 0.06 |
| No | 98/116 (84) | 8/13 (62) |  |
| Regular contact with sheep before start of FVMUU |  |  |  |
| Yes | 20/116 (17) | 5/13 (38) | 0.13 |
| No | 96/116 (83) | 8/13 (62) |  |
| Regular contact with poultry before start of FVMUU |  |  | 0.02 |
| Yes | 31/116 (27) | 8/13 (62) |  |
| No | 85/116 (73) | 5/13 (38) |  |
| Regular contact with horses before start of FVMUU |  |  |  |
| Yes | 65/116 (56) | 7/13 (54) | 1.00 |
| No | 51/116 (44) | 6/13 (46) |  |
| Regular contact with pigs before start of FVMUU |  |  |  |
| Yes | 13/116 (11) | 4/13 (31) | 0.07 |
| No | 103/116 (89) | 9/13 (69) |  |

Abbreviations: n=Number, N=Total number, BMI=Body Mass Index, FVMUU = Faculty of Veterinary
Medicine of Utrecht University, NA=Not available.
\* Two seronegative participants at baseline did not fill out a questionnaire.

Of the 118 seronegative participants at baseline, 78 started their study in 2006 and 40 in 2008 (Figure). Of those students, 23 seroconverted during the follow-up period of 362 person-years, which results in an incidence of 0.06 per person-year. None of the seroconverted participants reported that their general practitioner or medical specialist diagnosed them with acute Q fever. Additionally, none of the participants had a serological indication for a chronic infection.

**Figure.**
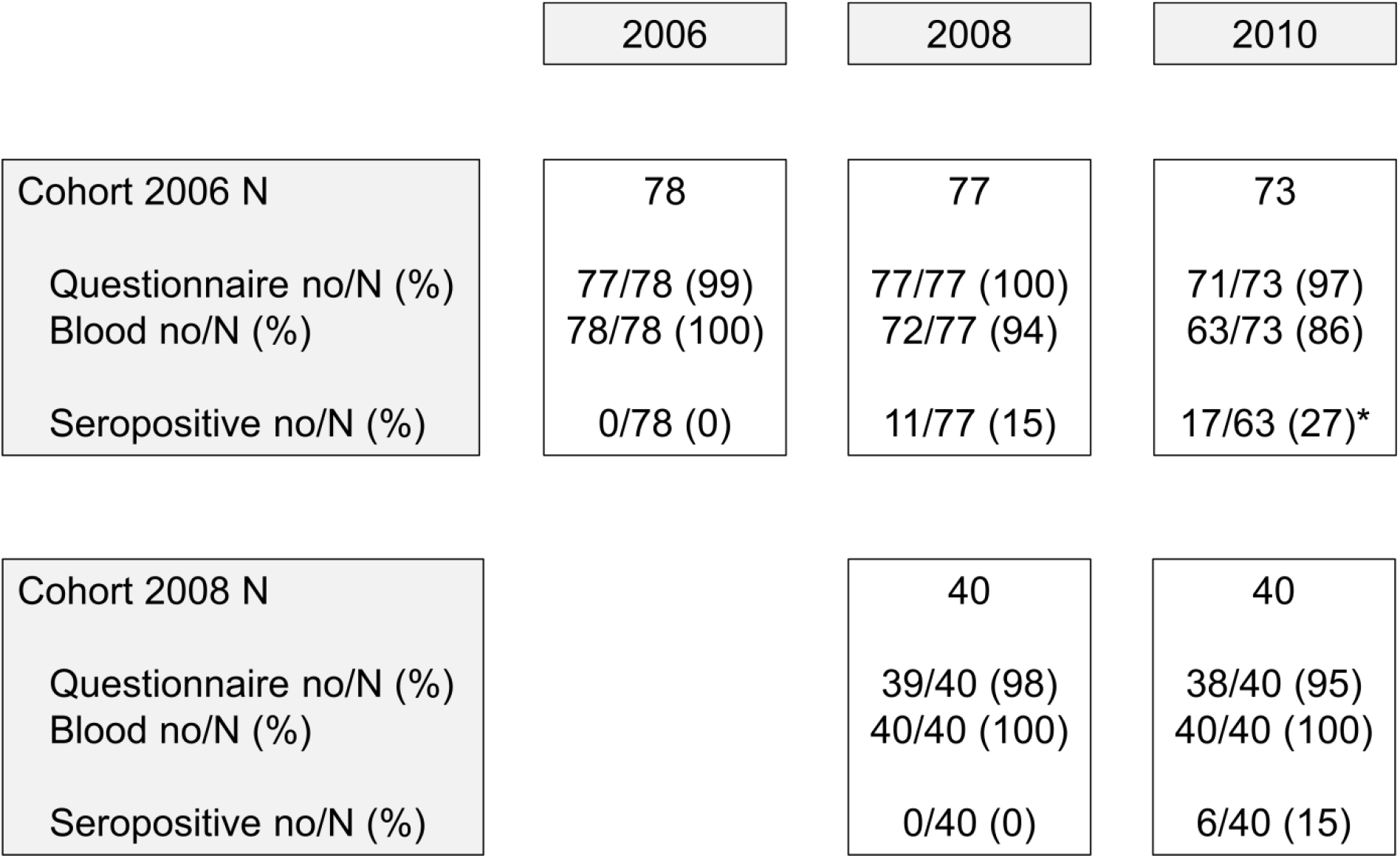
Follow-up timeline illustrating the number and percentages of seronegative participants at baseline, per follow-up moment. Abbreviations: no = the number in the corresponding group, N = total number of patients. * The 17 seropositive students in 2010 include the 11 seropositive students who already seroconverted between 2006 and 2008, who were censured from risk factor analysis in 2010.

Of the 20 investigated characteristics, “living on a sheep or goat farm”, “having contact with sheep outside the study”, and “working with hay, straw, silage grass or animal feed” during their study period outside the FVMUU increased the odds of seroconversion (p<0.05), in analyses adjusted for time since start study and cohort. Performing animal nursing on the farm where they lived tended to increase seroconversion (p=0.07) (Table 2).

**Table 2.** Characteristics from follow-up questionnaire in association with Q fever seroconversion among 118 veterinary students who were seronegative at start of their study in 2006 or 2008, the Netherlands

| Characteristic | OR (95% CI) | p value |
| --- | --- | --- |
| Age (years) |  |  |
| ≤ 20 | Reference |  |
| 21 | 0.9 (0.2-3.5) | 0.85 |
| ≥ 22 | 1.3 (0.4-4.2) | 0.69 |
| Gender |  |  |
| Male | Reference |  |
| Female | 0.7 (0.2-2.3) | 0.53 |
| Regularly exposed to cigarette smoke |  |  |
| Yes | 1.1 (0.4-2.8) | 0.81 |
| No | Reference |  |
| Living on a farm with cows |  |  |
| Yes | ND* |  |
| No |  |  |
| Living on a farm with sheep or goats |  |  |
| Yes | 6.2 (1.4-28.1) | 0.02 |
| No | Reference |  |
| Living on a farm with pigs |  |  |
| Yes | ND* |  |
| No |  |  |
| Living on a farm with chicken |  |  |
| Yes | 3.0 (0.3-35.0) | 0.39 |
| No | Reference |  |
| Having regular contact with cows outside study |  |  |
| Yes | 0.3 (0.1-2.7) | 0.31 |
| No | Reference |  |
| Having regular contact with goats outside study |  |  |
| Yes | 0.6 (0.1-3.8) | 0.56 |
| No | Reference |  |
| Having regular contact with horses outside study |  |  |
| Yes | 0.7 (0.3-1.7) | 0.40 |
| No | Reference |  |
| Having regular contact with pigs outside study |  |  |
| Yes | ND* |  |
| No |  |  |
| Having regular contact with chicken outside study |  |  |
| Yes | 0.5 (0.1-3.8) | 0.50 |
| No | Reference |  |
| Having regular contact with sheep outside study |  |  |
| Yes | 4.4 (1.2-16.7) | 0.03 |
| No | Reference |  |
| Performing animal nursing on the farm where you lived |  |  |
| Yes | 3.6 (0.9-14.3) | 0.07 |
| No | Reference |  |
| Working with straw/hay on the farm where you lived |  |  |
| Yes | 6.4 (1.6-26.1) | <0.01 |
| No | Reference |  |
| Working with fertilizers on the farm where you lived |  |  |
| Yes | 3.2 (0.5-19.6) | 0.21 |
| No | Reference |  |
| Performing plant nursing on the farm where you lived |  |  |
| Yes | 3.1 (0.3-33.5) | 0.35 |
| No | Reference |  |
| Number of years after start study† |  |  |
| Two | Reference |  |
| Four | 1.0 (0.3-2.9) | 0.96 |
| Cohort‡ |  |  |
| 2006 | Reference |  |
| 2008 | 0.7 (0.3-2.0) | 0.56 |
| Chosen specialization during study |  |  |
| Individually kept animals | Reference |  |
| Veterinary public health / farm animals | 1.6 (0.5-5.0) | 0.38 |

Abbreviations: CI=confidence interval, ND=Not determined, OR=Odds Ratio.
\*Not to be determined due to low numbers.
†Only adjusted for cohort.
‡Only adjusted for number of years after the study.

## Conclusion

In this longitudinal serological study among veterinary students, we found a *C. burnetii* incidence of 0.06 per person-year. None of the seroconverted participants self-reported that they were diagnosed with acute Q fever, suggesting all cases were mild or asymptomatic. Additionally, none of the participants had a serological indication for a chronic infection. In veterinarians, which is the studied group in an advanced stage of their career, a high IgG phase I seroprevalence was found. It remains debatable whether presence of antibodies in occupationally exposed people with frequent boosting is of clinical significance *(8)*.

Three non-study factors were associated with increased seroconversion. Proximity to (aborting) small ruminants, such as goats and sheep, has previously been reported as important risk factor in an outbreak in the Netherlands *(9)*. It is known that veterinary students have a high prevalence of animal contacts outside the study *(10)*. Next, contact with hay, straw, silage grass or animal feed, is a known risk factor for human Q fever *(11)*. A major acute Q fever outbreak occurred in the Netherlands from 2007 to 2010 *(12)*, as a result of which some students may also have contracted the infection. Although increased seroprevalence of Q fever in veterinary students prior to the outbreak has been reported *(3)*. We were not able to investigate study-related potential risk factors, such as followed courses due to two reasons. First, the curriculum changed during the investigation, so participants from the 2006 and 2008 cohort followed different courses, causing a low power in the analysis. Second, per cohort of students, there was little variation in followed courses.

Seroconversion incidence data is scarce. Of 246 seronegative culling workers 36 became IgG positive during the half year follow-up period, corresponding to an incidence of 0.29 per person-year *(13)*, which is higher than the incidence in veterinary students (0.06 per person-year).

A nationally funded Q fever vaccination program for occupationally exposed people was introduced in Australia in 2002 *(14)*. At this moment, the Health Council of the Netherlands does not advice to vaccinate occupationally exposed people *(15)*. The results of this study should be taken into account when reconsidering this advice.

In conclusion, we found a considerable *C. burnetii* seroconversion rate among veterinary students. Although the clinical significance of the presence of antibodies is unknown, it is advisable to inform students at the beginning of the study about the potential risks of acute and chronic Q fever.

## Data Availability

The data that support the findings of this study are available on request from the corresponding author.

## Acknowledgments

We would like to express our gratitude to all participants of the study. Many thanks to co-workers at the Institute for Risk Assessment Sciences involved in recruitment of the participants and serum sample collection (Lot Bannink, Manon Bogaerts, Isabella van Schothorst, Esmeralda Krop, Siegfried de Wind, Jack Spithoven, Marieke Oldenwening, Lidwien Smit, Haitske Gravenland and Bernadette Aalders) Next, we would like to thank all laboratory technicians of the serology unit of the Department of Medical Microbiology and Infection Control of the Jeroen Bosch Hospital for their work in analyzing serum samples. Last, we would like to thank Roel Coutinho of the Julius Center UMC Utrecht for critically reviewing the manuscript. The authors have no support or funding to report.

## Author Bio

Dr. de Lange is epidemiologist at the National Institute for Public Health and the Environment. She has conducted her PhD research on Q fever. Other research interests are respiratory infections, such as influenza and RSV.

## References

1. Maurin M, Raoult D. Q fever. Clin Microbiol Rev. 1999;12(4):518–53.

2. Van den Brom R, Schimmer B, Schneeberger PM, Swart WA, van der Hoek W, Vellema P. Seroepidemiological survey for Coxiella burnetii antibodies and associated risk factors in Dutch livestock veterinarians. PLoS One. 2013;8(1):e54021.

3. de Rooij MM, Schimmer B, Versteeg B, Schneeberger P, Berends BR, Heederik D, et al. Risk factors of Coxiella burnetii (Q fever) seropositivity in veterinary medicine students. PLoS One. 2012;7(2):e32108.

4. Meadows SL, Jones-Bitton A, McEwen SA, Jansen J, Patel SN, Filejski C, et al. Prevalence and risk factors for Coxiella burnetii seropositivity in small ruminant veterinarians and veterinary students in Ontario, Canada. Can Vet J. 2017;58(4):397–9.

5. Valencia MC, Rodriguez CO, Punet OG, de Blas Giral I. Q fever seroprevalence and associated risk factors among students from the Veterinary School of Zaragoza, Spain. Eur J Epidemiol. 2000;16(5):469–76.

6. Wegdam-Blans MC, Kampschreur LM, Delsing CE, Bleeker-Rovers CP, Sprong T, van Kasteren ME, et al. Chronic Q fever: review of the literature and a proposal of new diagnostic criteria. J Infect. 2012;64(3):247–59.

7. Liang K, Zeger SL. Longitudinal data analysis using generalized linear models. Biometrika. 1986;73(1):13–22.

8. Wielders CC, Boerman AW, Schimmer B, van den Brom R, Notermans DW, van der Hoek W, et al. Persistent high IgG phase I antibody levels against Coxiella burnetii among veterinarians compared to patients previously diagnosed with acute Q fever after three years of follow-up. PLoS One. 2015;10(1):e0116937.

9. Roest HI, Tilburg JJ, van der Hoek W, Vellema P, van Zijderveld FG, Klaassen CH, et al. The Q fever epidemic in The Netherlands: history, onset, response and reflection. Epidemiol Infect. 2011;139(1):1–12.

10. Samadi S, Spithoven J, Jamshidifard AR, Berends BR, Lipman L, Heederik DJ, et al. Allergy among veterinary medicine students in The Netherlands. Occup Environ Med. 2012;69(1):48–55.

11. Rustscheff S, Norlander L, Macellaro A, Sjostedt A, Vene S, Carlsson M. A case of Q fever acquired in Sweden and isolation of the probable ethiological agent, Coxiella burnetii from an indigenous source. Scand J Infect Dis. 2000;32(6):605–7.

12. van der Hoek W, Morroy G, Renders NH, Wever PC, Hermans MH, Leenders AC, et al. Epidemic Q fever in humans in the Netherlands. Adv Exp Med Biol. 2012;984:329–64.

13. Whelan J, Schimmer B, Schneeberger P, Meekelenkamp J, Ijff A, van der Hoek W, et al. Q fever among culling workers, the Netherlands, 2009-2010. Emerg Infect Dis. 2011;17(9):1719–23.

14. Gidding HF, Wallace C, Lawrence GL, McIntyre PB. Australia’s national Q fever vaccination program. Vaccine. 2009;27(14):2037–41.

15. Health Council of the Netherlands. Advisory letter employees and Q fever: criteria fo vaccination [in Ducth]. 2015. Report No.: 2015/07.

